# Comparison of Peripheral Nerve Injury Outcomes Between COVID-19 Survivors and Non-COVID Rehabilitation Inpatients: A Retrospective Study

**DOI:** 10.1101/2024.08.22.24312439

**Authors:** Antonio Mondríguez-González, Brian M. Rothemich, Manasi Sheth, Kevin N. Swong, Colin K. Franz

## Abstract

**Introduction:** Peripheral nerve injury (PNI) is associated with severe Coronavirus disease 2019 (COVID-19) survivorship. The diagnosis of PNI is often made by a physiatrist during a detailed functional assessment during an inpatient rehabilitation stay. COVID-19 patients have elevated rates of medical comorbidities, including risk factors for acquired PNI, such as diabetes mellitus and obesity. It is not known if the functional prognosis from PNI in COVID-19 survivors differs substantially from PNI in other inpatient rehabilitation populations.

**Objective:** To determine the prognosis of PNI associated with severe COVID-19 survivorship and compare it to PNI associated with other inpatient rehabilitation populations.

**Design:** Retrospective chart review study.

**Setting:** Single-center inpatient rehabilitation hospital in a large urban city.

**Patients:** Adult patients admitted to an inpatient rehabilitation hospital with PNI(s).

**Interventions:** Not applicable.

**Main Outcome Measures:** The primary outcome was the change in manual muscle testing (MMT) over time. Secondary outcomes included the rate of peripheral nerve surgery and the number of distinct PNI sites per patient.

**Results:** The analysis consisted of 60 subjects with PNI. We identified 30 subjects who had PNI associated with COVID-19 and were matched with 30 subjects with PNI not associated with COVID-19 who were diagnosed during their inpatient rehabilitation admission. The data collected included basic demographics, COVID-19 status immediately before inpatient rehabilitation admission, medical comorbidities, acute rehabilitation inpatient diagnosis, nerve injury location and mechanism of injury, muscles affected, and change in serial MMT, plus documentation of any surgical intervention. No significant difference was found between the improvement of MMT, surgery rate, or number of nerve injuries and COVID-19 status.

**Conclusion:** PNIs associated with severe COVID-19 survivorship have similar recovery patterns as those of other etiologies. This data is reassuring that PNI associated with COVID-19 may be managed similarly to other types of PNI.

## Introduction

Coronavirus disease 2019 (COVID-19) has profoundly affected the world’s health. As of August 2024, there have been 776 million confirmed disease cases.^1^ Furthermore, multiple studies suggest that survivorship issues from COVID-19 include an array of PNIs. Overall, although a variety of risk factors have been associated with acquired PNI from severe COVID-19, little is known about the clinical outcomes of these patients. To make matters worse, COVID-19 has been associated with multiple mononeuropathies in the same patient. For example, our group recently described a case series involving 66 PNIs found in 34 patients (average ∼2 PNI/patient).^2^ This association of PNI with COVID-19 survivorship was noted early in the pandemic, including an observation that 12/83 post-COVID-19 patients consecutively admitted for inpatient rehabilitation had an acquired PNI,^2^ and just a few weeks later, a separate group described mononeuritis multiplex patterns in 11/69 COVID-19 patients consecutively admitted to an intensive care unit (ICU).^3^ Irrespective to the COVID-19 pandemic, the rates of PNI worldwide are ill-defined. Still, the best estimates come from developed countries with national healthcare registries with rates between 11 to 14 cases per 100,000 population per year.^4,5^ This is likely an underestimation of PNI incidence. For example, PNI can be an under-reported complication in the perioperative setting,^6^ overlooked or missed in cases of complex polytrauma,^7^ and in many countries like the United States that lack a national registry to study PNI; there are higher rates of PNI from gun violence.^8^ Outcomes from PNI are known to be highly variable regardless of whether patients undergo nerve repair surgery.^8^ Therefore, it is often challenging to prognosticate patient outcomes accurately. Prior studies have implicated age, site of injury, type of injury, and extent of vascular damage as the most important factors.^8,9^

Given the large numbers of people who survived severe COVID-19 and consequently were exposed to this risk for acquiring a PNI, the question of whether or not PNI outcomes are worse in survivors of severe COVID-19 is both important and unanswered. The purpose of this retrospective chart review study is to begin to define neurological recovery patterns and prognosis of PNIs associated with severe COVID-19 versus other non-COVID-19 associations. Since we have access to a relatively large and chronic series of patients who acquired PNI in association with admission for acute COVID-19, we performed a retrospective study to compare their recovery patterns and outcomes in follow-up against those of non-COVID-19 associated PNI identified from inpatient rehabilitation patients diagnosed and treated in an overlapping time frame.

## Materials and Methods

### Study Subjects

Study approval was granted by the Northwestern University Institutional Review Board. A total of 60 subjects with a diagnosis of PNI were included in this study. PNI in the context of this study is defined as mononeuropathy caused by traumatic etiologies such as gunshots, puncture wounds, and high-impact collisions and nontraumatic etiologies such as entrapment/compression, critical illness, severe COVID-19 survivorship, or medications leading to an impact on muscle strength and motor function. Subjects were selected from a registry of known PNI cases treated at the Shirley Ryan AbilityLab (an academic freestanding inpatient rehabilitation facility) in Chicago, IL (USA). Furthermore, subjects included in this study were equal to or greater than 18 years of age, and their PNI(s) must have been supported by electrodiagnostic studies. All subjects were patients admitted to inpatient rehabilitation who had their initial electrodiagnostic study and MMT conducted upon their rehabilitation hospital admission prior to their PNI being diagnosed. Subjects were placed into the post-COVID-19 PNI group if they acquired PNI(s) after admission to an acute care hospital for severe COVID-19 pneumonia. The post-COVID-19 PNI group was admitted to inpatient rehabilitation due to generalized weakness and deconditioning as a result of severe COVID-19 survivorship and its associated hospital complications. Severe COVID-19 survivorship was defined as recovery from an acute care hospitalization for COVID-19 requiring the use of assistive respiratory devices. Patients who were under the age of 18 or did not fit the rest of the inclusion criteria were excluded from data collection.

### Study Outcomes

The primary outcome of this retrospective study is to compare the improvement of MMT from the initial diagnosis and after-treatment interventions.^10^ MMT was selected since it was well documented across patient charts and is a common clinical metric of peripheral nerve health. We were concerned that the improvement in MMT will be less in the post-COVID-19 group compared to other inpatient rehabilitation patients (control group) due to the severity of their preceding illness, thus indicating a slower or reduced recovery process. The secondary outcome is to compare the surgery rate between these two groups.

Due to the high rate of severe (axonotmesis) PNI associated with COVID-19,^2,^ we wondered if there may be a higher rate of nerve repair surgery within the post-COVID-19 group. The other secondary outcome is to compare the number of concomitant sites of PNI between the two groups. Since COVID-19 has been associated with mononeuritis multiplex patterns, we predicted that the post-COVID-19 group would be diagnosed with significantly more PNIs per patient.

### Data Collection

Data was collected by retrospectively reviewing patients’ electronic medical records with PNI from July 2018 to September 2023. A combination of electronic medical records from internal and external hospital systems was used to collect thorough data. We recorded several data points for each subject that consisted of age, race/ethnicity, whether or not the patient had COVID-19 before the PNI, comorbidities, acute rehabilitation inpatient status, mechanism of PNI, location of PNI, type of treatment intervention, electrodiagnostic studies at diagnosis, muscles affected by the PNI and their associated MMT scores at diagnosis and after treatment intervention. Each subject was screened for the following illnesses: diabetes mellitus, B12 deficiency, hypertension, hyperlipidemia, and obesity. The type of treatment was subdivided into no surgery, external neurolysis, direct nerve repair, nerve graft repair, and nerve transfer surgery. MMT grades were recorded with the standard 0-5 scale without modifiers (i.e., no 4+ or 4-). Only muscles tested during a documented physical exam and graded less than five were recorded at or within a month after initial electrodiagnostic studies. Follow-up MMT was done within 1 month of initial electrodiagnostic testing at either an outpatient or inpatient setting, depending on if the patient had been discharged from inpatient rehabilitation, although the latest available MMT was recorded for post-intervention data points within a maximum of two years of the start of the intervention (physical therapy, surgery).

### Data Preparation

When analyzing MMT data, the 60 subjects were further broken down into the specific nerve-injured and the associated muscle groups innervated by the nerve. Each nerve was then listed with its associated pre-intervention and post-intervention MMT grades. Each PNI was then categorized into one of three groups: no improvement, small improvement, and large improvement. No improvement was given if the difference in muscle grades was “0”. Small improvement was given if the muscle grade improved by one MMT grade, except for a “4,” progressing into a “5” to mitigate against the ceiling effect. Large improvement was given if the muscle grade improved to a “5” or improved by two or more MMT grades. Furthermore, if a PNI was not confirmed by a corresponding muscle group with a pre-intervention and post-intervention manual muscle test, it was removed from the data set (e.g., lateral femoral cutaneous nerve). In addition, if two or more muscle groups were associated with the PNI, only the muscle group with the highest level of improvement was included in the data set. If there was a tie in improvement level, then the most proximal muscle group with respect to the lesion site was included in the data set when paired with the PNI.

### Statistical Analysis

Following data collection and preparation, a Student’s t-test and several two-sided Fisher’s exact tests were appropriately used to determine the presence of a statistically significant difference between ages, sex, race/ethnicities, comorbidities, and mechanism of injury for the post-COVID-19 PNI versus other non-COVID-19 PNI. A p-value of 0.05 was used when testing the statistical significance.

A Mann-Whitney U test was used to evaluate whether improvement in strength grading from MMT (no improvement, small improvement, large improvement) significantly differs by the COVID-19 status of PNI (post-COVID-19 versus other non-COVID-19 associations). A p-value of 0.05 was used when testing the statistical significance.

A two-sided Fisher’s exact test was used to determine if there was a significant association between treatment intervention and the COVID-19 status of PNI. A 2x2 contingency table was made to record the number of observations for treatment intervention (surgery versus no surgery) and the COVID-19 status of PNI (post-COVID-19 versus other non-COVID-19 associations). A p-value of 0.05 was used when testing the statistical significance.

A two-sided Fisher’s exact test was used to determine if there was a significant association between the number of PNIs and post-COVID-19 status. A 2x2 contingency table was created to record the number of observations for each group. A p-value of 0.05 was used when testing the statistical significance.

### IRB and Data Storage

This research study was approved by the Northwestern University IRB before data collection. Since this project is a retrospective chart review with no contact with subjects, no informed consent or waiver of consent was needed to gather and conduct research. All data documents with patient identifiers were encrypted and only available to the study investigators. All data was initially entered into a password-protected RedCap project database without patient identifiers. Before statistical analysis, all patient identifiers were removed from the final data collection document.

## Results

### Demographics, Comorbidities, and Mechanism of Nerve Injury

A total of 60 subjects were analyzed in this study. Of these patients with PNIs, 30 subjects belong to the post-COVID-19 PNI group, while the remaining 30 subjects with other non-COVID-19 PNI belong to the control group. A summary of their characteristics can be seen in Table 1. Statistically significant differences were noted between the post-COVID-19 PNI group and the control group in the prevalence of diabetes (COVID n=19/30, non-COVID-19 3/30, p=0.0001), and hypertension (COVID n=22/30, non-COVID-19 8/30, p=0.0007). In addition, other statistically significant differences were found between the post-COVID-19 PNI group and the control group in the ages (COVID 55.9±12.2, non-COVID-19 43.2±16.2, p=0.001), and the Hispanic White racial/ethnicity group (COVID n=11/30, non-COVID-19 2/30, p=0.010). Lastly, the ratios of the mechanism of injury when comparing traumatic versus nontraumatic injuries proved to be statistically significant (COVID 0:30, non-COVID-19 19:11, p=0.0001). The ratios between sex, the remaining racial/ethnic groups, and obesity failed to reject the null hypothesis.

**Table 1.** Subject Characteristics. Significant p-values have been bolded.

| Subject Characteristics | COVID-19 | Non-COVID | p-values (sig<0.05) |
| --- | --- | --- | --- |
| Age | 55.9 ± 12.2 | 43.2 ± 16.2 | **p=0.001** |
| Sex M:F | 26:4 (n=30) | 21:9 (n=30) | p=0.209 |
| Race/Ethnicities | Non-Hispanic White<br>33.3% (10/30) | Non-Hispanic White<br>60.0% (18/30) | Non-Hispanic White:<br>p=0.069 |
|  | Non-Hispanic<br>Black/African<br>American 23.3% (7/30) | Non-Hispanic<br>Black/African<br>American 16.7% (5/30) | Non-Hispanic<br>Black/African<br>American: p=0.748 |
|  | Hispanic White 36.7%<br>(11/30) | Hispanic White 6.7%<br>(2/30) | Hispanic White:<br>**p=0.010** |
|  | Unknown 6.7% (2/30) | Unknown 16.7% (5/30) | Unknown: p=0.437 |
| Comorbidities | Diabetes 63.3% (19/30) | Diabetes 10.0% (3/30) | Diabetes: **p=0.0001** |
|  | Obesity 43.3% (13/30) | Obesity 30.0% (9/30) | Obesity: p=0.422 |
|  | Hypertension 73.3%<br>(22/30) | Hypertension 26.7%<br>(8/30) | Hypertension:<br>**p=0.0007** |
| Mechanism of Injury<br>Traumatic:Nontraumatic | 0:30 (n=30) | 19:11 (n=30) | **p=0.0001** |

**Table 2.**
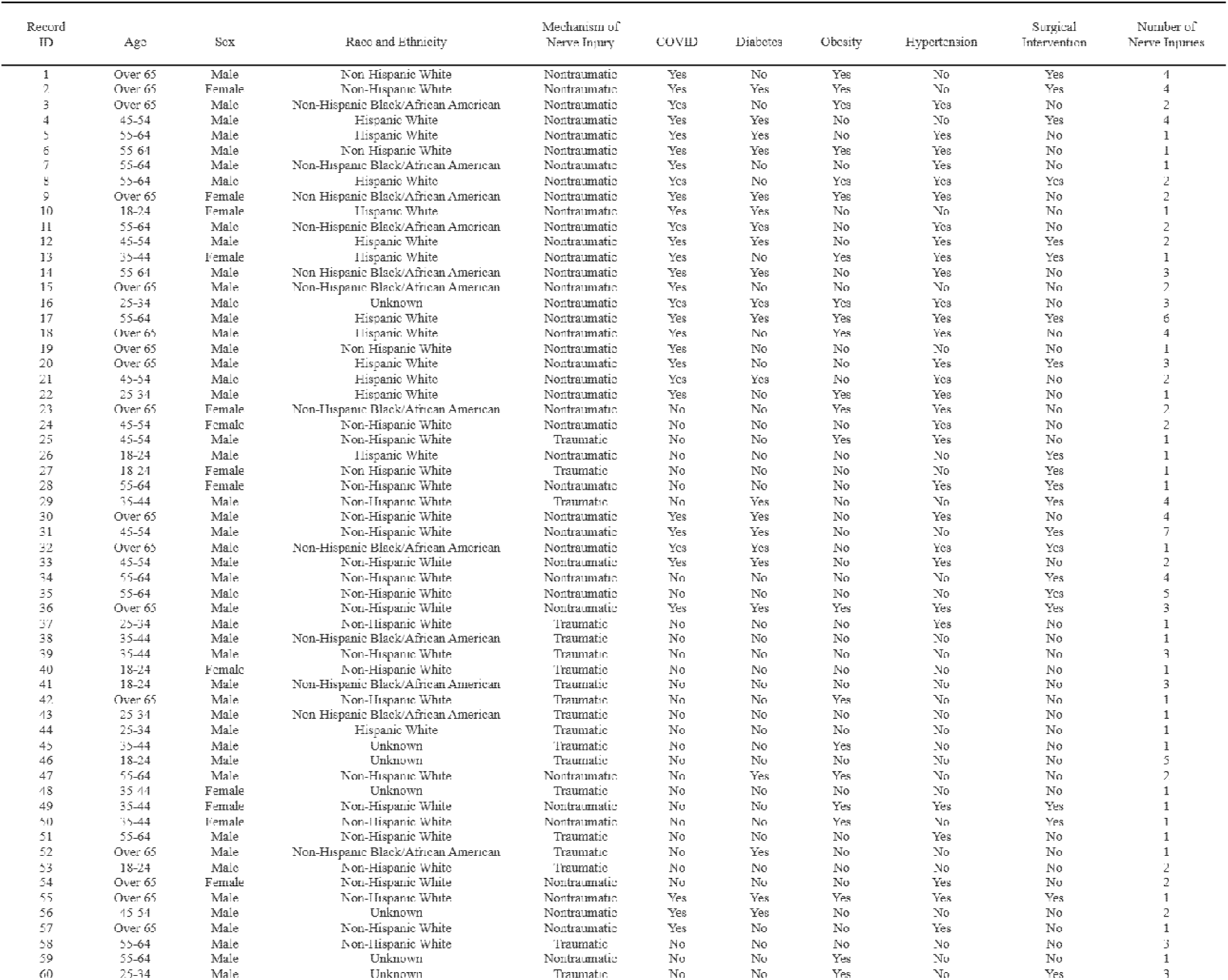
Demographics, Comorbidities, and Nerve Injuries.

**Table 3.**
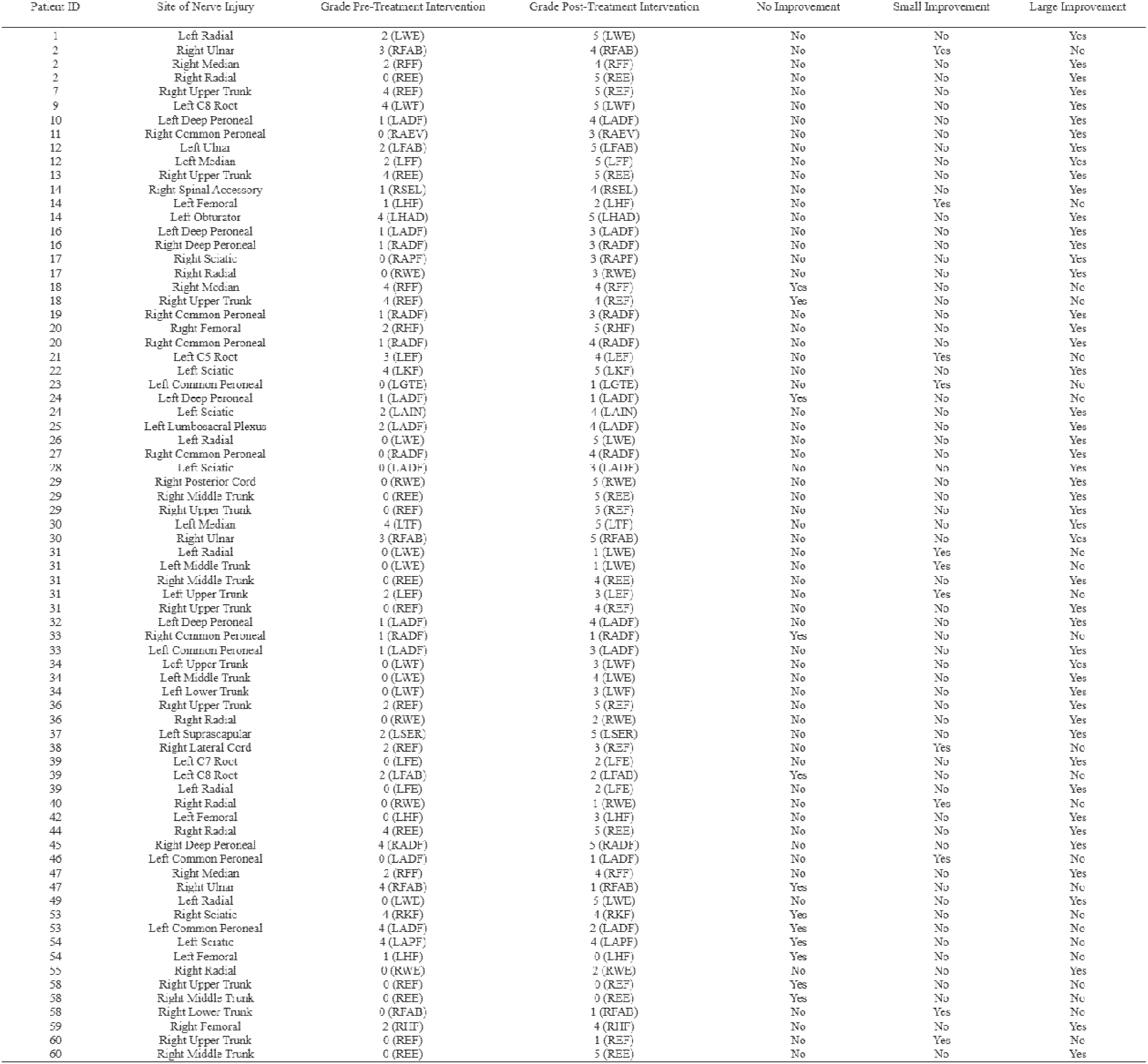
Manual Muscle Testing. Small Improvement (increase by one grade), Large Improvement (Increase to 5 or increase by two or more grades); (LSEL)/(RSEL)-left/right shoulder elevators, (LEF)/(REF)-left/right elbow flexors, (LEE)/(REE)-left/right elbow extensors, (LWF)/(RWF)-left/right wrist flexors, (LWE)/(RWE)-left/right wrist extensors, (LFF)/(RFF)- left/right finger flexors, (LFE)/(RFE)-left/right finger extensors, (LFAB)/(RFAB)-left/right finger abductors, (LTF)/(RTF)-left/right thumb flexors, (LHF)/(RHF)-left/right hip flexors, (LHAD)/(RHAD)-left/right hip adductors, (LKF)/(RKF)-left/right knee flexors, (LADF)/(RADF)-left/right ankle dorsiflexors, (LAPF)/(RAPF)-left/right ankle plantarflexors, (LAEV)/(RAEV)-left/right ankle evertors, (LGTE)/(RGTE)-left/right great toe extensors

### MMT Comparison Results

Improvements in MMT data are summarized in Figure 1A. A Mann-Whitney U test was used to evaluate whether improvement in strength grading from MMT (no improvement, small improvement, large improvement) significantly differs by the COVID-19 status of PNI (post-COVID-19 versus other non-COVID-19 associations). The null hypothesis stated no significant difference exists between the improvements in manual muscle tests and the COVID-19 status of PNI. The results indicated no significant difference between the improvement in manual muscle tests and the COVID-19 status of PNI (z = 1.5195, p = 0.1285).

**Figure 1.**
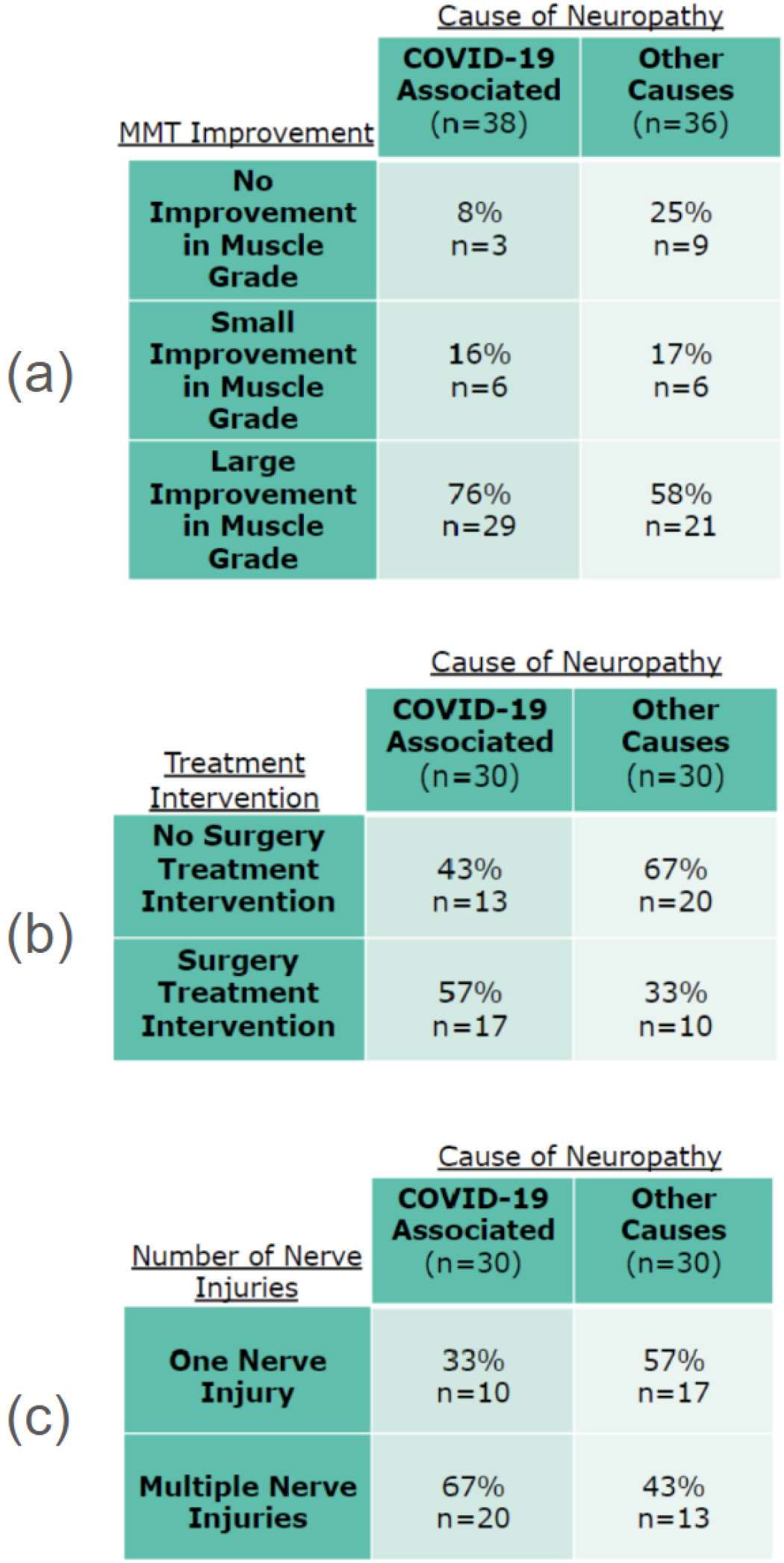
Contingency Tables. (a) MMT Improvement. Small Improvement: increase by one grade; Large Improvement: increase to 5/5 or increase by two or more grades. (b) Number of Surgeries. (c) Number of Nerve Injuries.

### Surgery Comparison Results

Surgical intervention data for each subject’s PNI(s) is displayed in Table 1 and summarized in Figure 1B. We predicted that there would be a significantly higher surgery rate within the post-COVID-19 group, thus indicating that they require a more complicated recovery process due to the severity of the injuries. A Fisher’s exact test was conducted to test the prediction that a significant association exists between treatment intervention and the COVID-19 status of PNI. The null hypothesis stated that no significant association exists between treatment intervention and the COVID-19 status of PNI. Using a 0.05 alpha, the null hypothesis failed to be rejected (two-tailed p = 0.1188). Therefore, there is no significant association between treatment intervention (no surgery versus surgery) and the COVID-19 status of PNI (post-COVID-19 versus other non-COVID-19 associations).

### Number of Nerve Injuries Comparison Results

The number of nerve injuries for each subject is shown in Table 1 and summarized in Figure 1C. We predicted that there would be a significantly higher number of PNIs within the post-COVID-19 group. A Fisher’s exact test was conducted as outlined in the Materials and Methods section to test this prediction that there is a significant association between the number of PNIs and the COVID-19 status of the PNI. The null hypothesis stated that no significant association exists between the number of PNIs and the COVID-19 status of the PNI. Using a 0.05 alpha, the null hypothesis failed to be rejected (two-tailed p = 0.1188). Therefore, there is no significant association between the number of PNIs (one injury versus multiple injuries) and the COVID-19 status of the PNI (post-COVID-19 versus other non-COVID-19 associations).

## Discussion

Severe COVID-19 survivorship is associated with PNIs. Many of these COVID-19-associated PNIs tend to be severe, primarily associated with axonal injuries, and even with proper medical care, lead to significant disability in many patients.^2^ This prior data implied that COVID-19-associated PNIs may have less favorable outcomes than other types of PNIs. Upon initial evaluation, the COVID-19 PNI group had a statistically significant older sample and a higher presence of comorbidities such as diabetes and hypertension, as seen in Table 1. Older age, diabetes, and hypertension are risk factors for developing PNIs, while older age is a risk factor for poorer PNI prognosis.^9^ However, our retrospective analysis revealed no significant associations between improvement in MMT and whether or not the PNI was associated with COVID-19. Our findings reassure us that COVID-19-associated PNI patients do not have substantially different outcomes. In addition, there were no differences in the need for surgical intervention or the number of PNI sites/patients and COVID-19. Together, these results indicate COVID-19 COVID-19-associated PNI patients do not appear to have inherently worse outcomes than non-COVID-19 PNI patients. These results support using existing treatment guidelines for PNIs in this specific patient population.

Severe COVID-19 patients appear to have an increased propensity for acquired PNI, and the potential reasons for this are likely multifactorial. Although the data is limited, in one neuropathological study of patients who died from COVID-19, muscle and nerve tissues exhibited signs of immune-mediated damage without direct infection by SARS-CoV-2, underscoring the impact of immune response rather than viral invasion in nerve injuries associated with the disease.^11^ It appears that an interplay of inflammatory and other immune-mediated factors results in a cytokine storm that directly damages nerve tissue and supporting vasculature,^12,13,14,15^ combined with mechanical loading of nerves due to patient positioning while admitted in the ICU.^16,17,18^ Indeed, our group previously noted that PNI localizations seemed heavily biased to sites of known vulnerability to mechanical loading from direct compression or traction mechanisms.^2^

PNI management can vary depending on whether they are open or closed injuries. Open injuries from traumatic mechanisms such as laceration or transection generally benefit from early surgical repair (re-anastomosis, grafting, etc.) when they are identified acutely. Meanwhile, closed injuries can be due to more diverse etiologies, from non-penetrating traumatic injury to chronic nerve entrapment injury, such as carpal tunnel syndrome.^19^ Generally speaking, COVID-19-associated PNIs are closed injuries. Closed injuries require a thorough clinical assessment, usually including electrodiagnostic studies and serial reassessments, which can include advanced imaging with repeat electrodiagnostic studies and physical examination. If a closed PNI shows no signs of spontaneous axon regeneration, then patients may consider whether or not to undergo surgery, which may involve repair methods such as neurolysis, neuroma resection, and/or graft repair, as well as nerve transfer surgery.^19^ Physical and/or occupational therapy is an essential part of the pre-habilitation and post-surgical rehabilitation process. Interestingly, despite the non-COVID-19 PNI group of our study containing many more traumatic mechanisms of injury, the rate of surgery used to manage these two patient groups was not statistically different. The significance of this finding is uncertain, given the retrospective nature of our study.

Since the recovery of COVID-19 patients with PNIs does not differ substantially from the control group, it appears that similar treatment algorithms may be applied to both groups. In addition to reinforcing current guidelines on PNI treatment management for severe COVID-19 survivors, this study should help rehabilitation physicians evaluate the prognosis of post-COVID-19 patients with PNIs. This study’s findings suggest no difference in muscle strength recovery regardless of the mechanism of injury and association with severe COVID-19 survivorship. Ninety-two percent of patients with COVID-19-associated PNI gained back some level of muscle improvement, while 75% of other causes of PNI improved in motor deficits. A majority of patients experiencing motor improvement after PNI treatments is not unexpected and is in line with multiple prior studies. For example, Kim et al. demonstrate that 73% of patients with thoracic outlet syndrome-associated motor deficits improved after appropriate treatment.^8^ Furthermore, another study with 107 patients with a variety of upper limb PNIs had a statistically significant improvement in all 107 patients’ muscle strength after appropriate treatment.^20^ Despite state-of-the-art treatments being made available to all PNI patients in this study, functional recoveries were near-universally incomplete.^2,8^ Partial functional recovery of PNIs remains a significant burden of disability of post-ICU survivors, including those that develop PNIs after severe COVID-19 survivorship.^21^ Although these PNIs associated with COVID-19 remain primarily axonal,^2^ no FDA-approved treatments are available to improve axon regeneration in this patient population. More clinical research should be conducted to find effective axonal regeneration treatments to reduce the functional disability of these patients.

Evaluation of the study samples revealed possible health disparities due to the overrepresentation of a socially disadvantaged population. In the COVID-19-associated PNI sample, the distribution of race/ethnicity was 33.3%, 23.3%, and 36.7% for non-Hispanic white, non-Hispanic Black/African American, and Hispanic white, respectively. According to the 2018-2022 United States Census, Chicago’s distribution of race/ethnicity was 32.7%, 28.8%, and 29.0% for non-Hispanic white, non-Hispanic Black/African American, and Hispanic white, respectively.^22^ The data demonstrates a large overrepresentation of patients who identify as Hispanic white in the severe COVID-19 PNI group. As seen in Table 1, a statistical analysis confirmed an overrepresentation of people who identify as Hispanic white in the COVID-19 PNI group. This overrepresentation could be due to a variety of factors. The Hispanic population in Chicago was the most likely ethnic group to work in essential and high-exposure jobs during the COVID-19 pandemic.^23^ In addition, there are social and economic factors within this ethnic group that reduce their access to healthcare, thus increasing their chances of developing complications due to severe COVID-19 survivorship.^24^ These risk factors and health disparities affiliated with COVID-19-associated PNI can be further investigated in a subsequent study.

It is important to recognize the scope of clinical impact this study has regarding the increased prevalence of PNIs in the study population. The rate of focal neuropathies associated with severe COVID-19 survivorship at inpatient rehabilitation facilities is approximately 10% compared to approximately 0.5% in general rehab patients over the same time period.^2^ PNIs subject patients to a substantial amount of long-term disability due to sensorimotor deficits and chronic neuropathic pain. Considering the high prevalence of PNIs associated with COVID-19 in the inpatient rehabilitation setting, physicians can implement the results from this study by confirming the effectiveness of their own clinical practices and existing treatment guidelines for addressing PNIs.

## Limitations

The limitations need to be addressed before considering the full implications of this study. There is a relatively small sample of 60 subjects used in this study due to the limited number of qualifying subjects from a single healthcare facility. Both the smaller sample size and the location where the sample was collected reduce external validity; however, this study is still the largest single data set to report COVID-19 PNI long-term outcomes. In addition, since patient follow-ups occurred inconsistently and at varied time frames, the post-treatment MMT data was collected at different time intervals among the patients. Selection bias resulted from several patients or multiple muscle groups not being included in the MMT data analysis due to incomplete follow-up data. Incomplete follow-up data either resulted from patients not returning for follow-up visits entirely or follow-up visits only containing partial MMT data compared to their initial visit. These issues could potentially introduce bias by disproportionately increasing or decreasing the improvement in MMT for one or both groups. Lastly, due to the retrospective design of this study, outcome data for muscle strength improvement was limited to an outcome measure that is used routinely in clinical practice, such as MMT, rather than more objective and continuous data, such as torque production or electrophysical recordings.

### Future Works

Several of these limitations can be addressed in future extensions of this study. For example, a larger sample size could increase the power of the results. The inclusion of multiple hospital systems, such as collaboration with other hospitals within our geographical region, could increase the external validity of the results. Furthermore, we hope to include phrenic neuropathy data in future iterations of this study by tracking improvements in diaphragm thickness ratios. Lastly, a multivariable regression analysis might reveal complex associations not outlined in this paper, such as manual muscle improvement associated with race, gender, number of nerve injuries, treatment intervention, and COVID-19 status of nerve injury.

## Conclusion

This data is impactful to the medical community as there is no literature to our knowledge that outlines the prognosis of this particular study population. This research fills a gap in the literature by providing a better understanding of the prognosis of post-COVID-19 PNIs. Because the results suggest there is no significant difference in prognosis between PNIs and their COVID-19 status, it can be inferred that physicians should not treat the two groups differently.

## Data Availability

All data produced in the present study are available upon reasonable request to the authors

