## Supplementary figures and images for "Comparison of Peripheral Nerve Injury Outcomes Between COVID-19 Survivors and Non-COVID Rehabilitation Inpatients: A Retrospective Study"

### Supplemental Table 2

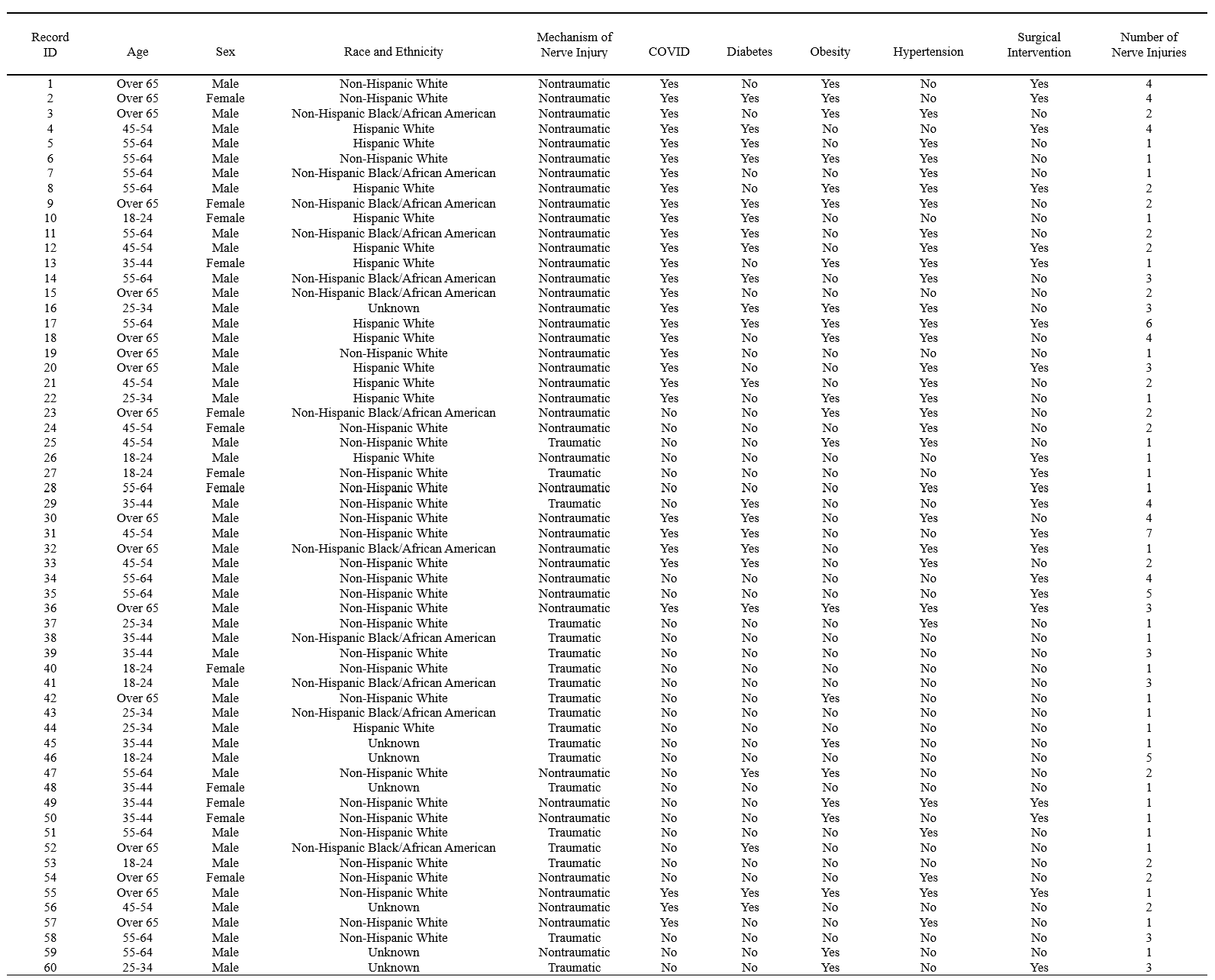

**Table 2. Demographics, Comorbidities, and Nerve Injuries.**
