## Supplemental Table 3 for "Comparison of Peripheral Nerve Injury Outcomes Between COVID-19 Survivors and Non-COVID Rehabilitation Inpatients: A Retrospective Study"

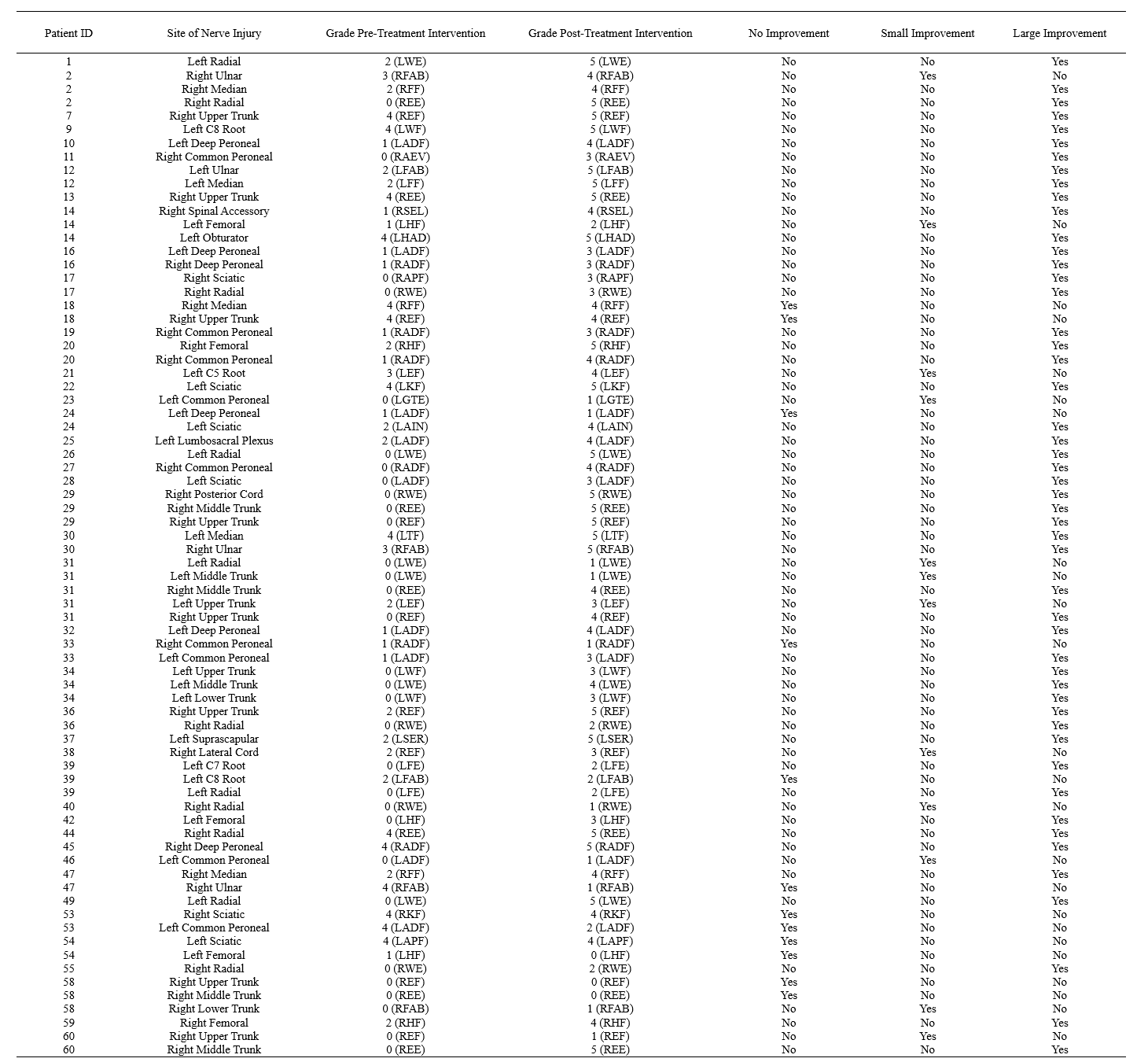
**Table 3. Manual Muscle Testing.** Small Improvement (increase by one grade), Large Improvement (Increase to 5 or increase by two or more grades); (LSEL)/(RSEL)- left/right shoulder elevators, (LEF)/(REF)- left/right elbow flexors, (LEE)/(REE)- left/right elbow extensors, (LWF)/(RWF)- left/right wrist flexors, (LWE)/(RWE)- left/right wrist extensors, (LFF)/(RFF)- left/right finger flexors, (LFE)/(RFE)- left/right finger extensors, (LFAB)/(RFAB)- left/right finger abductors, (LTF)/(RTF)- left/right thumb flexors, (LHF)/(RHF)- left/right hip flexors, (LHAD)/(RHAD)- left/right hip adductors, (LKF)/(RKF)- left/right knee flexors, (LADF)/(RADF)- left/right ankle dorsiflexors, (LAPF)/(RAPF)- left/right ankle plantarflexors, (LAEV)/(RAEV)- left/right ankle evertors, (LGTE)/(RGTE)- left/right great toe extensors
